# COVID-19 Impact on Patients with Immune-Mediated Rheumatic Disease: A Comparative Study of Disease Activity and Psychological Well-Being Over Six Months

**DOI:** 10.1101/2024.03.18.24304464

**Authors:** Claudia Marques, Marcelo M Pinheiro, Jennifer Lopes, Sandra Lúcia Euzébio Ribeiro, Mary Vânia Marinho de Castro, Lilian David de Azevedo Valadares, Aline Ranzolin, Nicole Pamplona Bueno de Andrade, Rafaela Cavalheiro do Espírito Santo, Nafice Costa Araújo, Cintya Martins Vieira, Valéria Valim, Flavia Patricia Sena Teixeira Santos, Laurindo Ferreira da Rocha Junior, Adriana Maria Kakehasi, Ana Paula Monteiro Gomides Reis, Edgard Torres dos Reis-Neto, Gecilmara Salviato Pileggi, Gilda Aparecida Ferreira, Licia Maria Henrique da Mota, Ricardo Machado Xavier

## Abstract

**Objectives:** To compare the impact of COVID-19 on clinical status and psychological condition in patients with immune-mediated rheumatic diseases (IMRD) infected by SARS-CoV-2 with IMRD controls not infected, during a 6-month follow-up.

**Methods:** The ReumaCoV Brasil is a longitudinal study designed to follow-up IMRD patients for 6 months after COVID-19 (cases) compared with IMRD patients no COVID-19 (controls). Clinical data, disease activity measurements and current treatment regarding IMRD, and COVID-19 outcomes were evaluated in all patients. Disease activity was assessed through validated tools at inclusion and at 3 and 6 months post-COVID-19. The FACIT<u>-F</u> (Functional Assessment of Chronic Illness Therapy) and DASS 21 (Depression, Anxiety and Stress Scale – 21 Items) questionnaires were also applied at 6 months after COVID-19 in both groups before large-scale vaccination. The significance level was set as p<0.05, with a 95% confidence interval.

**Results:** A total of 601 patients were evaluated, being 321 cases (IMRD COVID-19+) and 280 controls (IMRD COVID-19 –), predominantly female with similar median age. No significant differences were noted in demographic data between the groups, including comorbidities, disease duration, and IMRD. Disease activity assessment over a 6-month follow-up showed no significant difference between cases and controls. While mean activity scores did not differ significantly, some patients reported worsened disease activity post-COVID-19, particularly in rheumatoid arthritis (RA) (32.2%) and systemic lupus erythematosus (SLE) (23.3%). Post-COVID-19 worsening in RA patients correlated with medical global assessment (MGA) and CDAI scores, with a moderate to large effect size. Diabetes mellitus showed a positive association (OR=7.15), while TNF inhibitors showed a protective effect (OR=0.51). Comparing SLEDAI pre– and post-COVID-19, a minority showed increased scores, with few requiring treatment changes. Fatigue, depression, anxiety, and stress were significantly higher in cases compared to controls. Worsening disease activity post-COVID correlated with worsened FACIT-F and DASS-21 stress scale in RA patients. No significant associations were found between COVID-19 outcomes and post-COVID-19 disease activity or psychological assessments.

**Conclusions:** Post-COVID-19 IMRD patients show significant psychological well-being deterioration despite similar disease activity scores. The variability in reports on IMRD flares and the potential trigger of SARS-CoV-2 for autoimmune manifestations underline the need for detailed clinical assessment and a comprehensive approach to managing them.

## Introduction

The COVID-19 pandemic caused by the SARS-CoV-2 virus represents one of the greatest public health challenges worldwide. The infection has raised concerns about how the disease behaves in patients with immune-mediated rheumatic diseases (IMRD), particularly in terms of flares after infection and immunosuppressive treatment as potential risk factors for COVID-19 severity. Since 2020, much information has been published about the understanding of this association and its implications, but few studies have evaluated rheumatic disease flare prospectively post-COVID-19 (1) and existing data are still controversial.

Some individuals with immune-mediated rheumatic diseases (IMRD) may experience exacerbation following SARS-CoV-2 infection. However, uncertainties persist regarding whether these exacerbations are directly linked to disease activity or represent manifestations associated with COVID-19 itself. The phenomenon commonly referred to as “Long COVID” has been associated with a diverse range of clinical manifestations, encompassing joint pain, fatigue, and psychological symptoms like depression and anxiety, which can endure for several months post-infection [2]. In patients with IMRD, the presence of these symptoms may be connected to disease flares, introducing ambiguity in determining the appropriate course of treatment. Additionally, some authors have highlighted psychosomatic aspects or a fibromyalgia-like presentation, particularly as the prevalence increased following the initial wave of the pandemic. This increase appears to be more correlated with heightened fears, uncertainties, and lockdown strategies, as biological abnormalities such as impaired immune response or viral persistence have not been conclusively identified yet (2).

This study aimed to assess the disease activity status in patients with immune-mediated rheumatic diseases (IMRD) who had experienced COVID-19, comparing them to those who did not contract the virus, over a 6-month follow-up period. Furthermore, the objective was to delineate the prevalence of fatigue and psychological disorders, including depressive symptoms, anxiety, and stress, and to investigate the potential association of these symptoms with IMRD flares.

## Materials and Methods

### Study design – ReumaCoV Brasil Cohort

This paper presents the protocol for the ReumaCoV-Brasil Registry protocol as described elsewhere (3). Briefly, a prospective observational cohort was carried out in 13 university centers distributed along all five Brazilian geographic regions, including patients with IMRD and COVID-19 and a comparison group (patients with only IMRD), with a follow-up time of 6 months. Patients were evaluated in three consecutive visits: visit 1 (inclusion), visit 2 (3 months) and visit 3 (6 months), from May to December 2020. Disease activity status in the medical record of the most recent consultation, carried out at least in the last 6 months before inclusion, was considered as pre-COVID status. The post-COVID status was that which the patient presented at inclusion in the study.

### Disease activity evaluation

In all visits, data related to disease activity was collected, using specific validated instrument, mentioned here are those included in this analysis: Clinical Disease Activity Index (CDAI)(4) for RA patients, Systemic Lupus Erythematosus Disease Activity Index 2000 (SLEDAI-2k) (5)in SLE patients, Bath Ankylosing Spondylitis Disease Activity Index (BASDAI)(6) for patients with SpA. Besides the activity scores, medical global assessment (MGA) and patient global assessment (PGA) were collected regarding disease activity, using a visual analogue scale (VAS) from 0 to 10 points, being 10 the better score, and the patient opinion regarding DRIM worst since the last visit, also using a VAS from 0 to 10.

### Disease flare definition

– Rheumatoid arthritis: worsening of symptoms compared to the pre-COVID-19 state plus an increase of 4.5 points in the CDAI and/or need to change treatment (7).
– Systemic Lupus Erythematosus: worsening of symptoms compared to the pre-COVID-19 state plus an increase at least 4 points in the SLEDAI and/or need to change treatment (8).
– Spondyloarthritis: worsening of symptoms compared to the pre-COVID-19 state plus an increase at least 2 points in BASDAI and/or need to change treatment(9).

### Psychological distress evaluation

At visit 3, two questionnaires were administered: one to assess fatigue, using The Functional Assessment of Chronic Illness Therapy – Fatigue Scale (FACIT-F)(10), and other to assess mental symptoms, using The Depression, Anxiety and Stress Scale (DASS-21)(11).

The FACIT-F Scale is a short, 13-item tool that measures an individual’s level of fatigue during their usual daily activities over the past week. The level of fatigue is measured on a four-point Likert scale (4 = not at all fatigued to 0 = very much fatigued). The score varies from 0 to 52, and the lower the result, the higher the level of fatigue.

DASS-21 is used as a quantitative measure of distress along the 3 axes of depression (D), anxiety (A) and stress (S) reactions and management. Each of the questions is rated from 0 to 3. Therefore, each of the axes presents partial scores of 0 to 18-24 depending on the number of questions assigned.

The study was registered at Brazilian Registry of Clinical Trials RBR-33YTQC and approved by the Brazilian National Research Council (CONEP) under number 3.955.206. All participants were requested to provide explicit opt-in consent prior to participating in the survey.

### Statical analysis

The Kolmogorov Smirnoff test was performed to verify the normal distribution of continuous variables. As non-normality was verified, the median was used as a measure of central tendency with the respective interquartile range (IQR).

To verify the association among categorical or continuous variables, the chi-square or Fisher test and Pearson test were used, respectively. The Mann-Whitney test was used to compare means and to estimate the effect size by calculating the r index and the R squared (R^2^). The r index was used to evaluate the correlation between two variables: small effect size: 0.10; medium effect size: 0.30 and large effect size: 0.50.

To determine the effect size (ES) for comparing means, Cohen’s d test was used, interpreted as follows: 0.2 to 0.3 for small effect (the difference between the groups is subtle); around 0.5 as medium effect (moderate difference between the groups) and 0.8 or more as large effect (substantial difference between the groups). To compare the variables across three visits, the One-way Anova test (CI 95%) was used for repeated measures.

For this analysis, the SPSS statistical package, version 29.0.2.0 and the Graph Pad Prism software, Version 10.1.1 (270), November 21, 2023, were used. The significance level was set at p < 0.05, with 95% confidence interval.

## Results

A total of 601 patients were evaluated, including 321 cases (IMRD COVID+) and 280 controls (IMRD COVID-), with the majority being female with similar median age. No differences were observed regarding demographic data between the two groups, including comorbidities, disease duration and IMRD. A higher frequency of social isolation was observed in the control group (p=0.001), as well as they were on TNFi treatment (p=0.003). Table 1 summarizes the clinical and demographic data at baseline.

**Table 1.** Clinical and demographic data of the sample, comparing IMRD patients with COVID-19 versus without COVID-19 at baseline. Chi square test, unless otherwise noted.

|  | Cases<br>(IMRD COVID +)<br>n=321 |  | Controls<br>(IMRD COVID -)<br>N=280 |  |  |
| --- | --- | --- | --- | --- | --- |
|  | n | % | n | % | P-value |
| **Female** | 275 | 85.7 | 237 | 84.6 | 0.724 |
| **Age (median, IQR)** | 46 (37-57) |  | 49 (38-57) |  | 0.136** |
| **Ethnicity** |  |  |  |  |  |
| *White* | 105 | 32.7 | 99 | 35.3 | 0.601 |
| *Non-white* | 216 | 67.3 | 181 | 64.7 |  |
| **Instruction** |  |  |  |  |  |
| *Illiterate* | 1 | 0.3 | 2 | 0.7 | 0.631 |
| *< 8 years* | 78 | 24.3 | 74 | 26.5 |  |
| *> 8 years* | 242 | 75.4 | 204 | 72.8 |  |
| **Profession** |  |  |  |  |  |
| *Public attendance* | 41 | 12.8 | 33 | 11.7 | **0.030** |
| *Health* | 34 | 10.6 | 13 | 4.7 |  |
| *Security* | 8 | 2.5 | 3 | 1.1 |  |
| *Education* | 32 | 10.0 | 25 | 9.0 |  |
| *Housewife* | 90 | 28.1 | 77 | 27.6 |  |
| *Others* | 116 | 36.0 | 129 | 45.9 |  |
| **Active at work** | 166 | 51.9 | 125 | 44.8 | 0.084 |
| **Social isolation** | 162 | 50.5 | 188 | 67.1 | **0.001** |
| **Comorbidities** |  |  |  |  |  |
| *Hypertension* | 123 | 38.3 | 120 | 42.9 | 0.258 |
| *Obesity* | 48 | 15.0 | 34 | 12.1 | 0.317 |
| *Diabetes* | 31 | 9.7 | 22 | 7.9 | 0.438 |
| *Lung disease* | 20 | 6.2 | 16 | 5.7 | 0.790 |
| *Kidney disease* | 20 | 6.2 | 16 | 5.7 | 0.790 |
| *Cardiopathy* | 12 | 4.3 | 13 | 4.6 | 0.734 |
| *No comorbidities* | 107 | 33.3 | 99 | 35.4 | 0.602 |
| **Smoking** | 11 | 3.7 | 15 | 4.6 | 0.580 |
| **Ethilism** | 10 | 3.1 | 8 | 2.8 | 0.835 |
| **Diagnosis** |  |  |  |  |  |
| *SLE* | 137 | 42.5 | 112 | 39.4 | 0.183 |
| *RA* | 93 | 28.9 | 105 | 37.0 |  |
| *Axial SpA* | 23 | 7.1 | 30 | 10.6 |  |
| *Psoriatic arthritis* | 21 | 6.5 | 12 | 36.4 |  |
| *Systemic sclerosis* | 17 | 5.3 | 10 | 3.6 |  |
| *Sjögren disease* | 12 | 3.7 | 5 | 1.8 |  |
| *Miopathy* | 7 | 2.2 | 2 | 0.7 |  |
| *Others* | 11 | 3.4 | 4 | 1.4 |  |
| **Disease duration, years (median, IQR)** | 10 (5-15) |  | 10 (6-18) |  | 0.146** |
| **Medication IMRD** |  |  |  |  |  |
| *Hydroxychloroquine* | 139 | 43.3 | 113 | 40.4 | 0.465 |
| *Oral corticosteroid* | 137 | 42.7 | 109 | 38.9 | 0.351 |
| *Methotrexate* | 72 | 22.4 | 73 | 26.1 | 0.298 |
| *TNFi* | 61 | 19.0 | 82 | 29.3 | **0.003** |
| *Azathioprine* | 48 | 15.0 | 30 | 10.7 | 0.123 |
| *Leflunomide* | 36 | 11.2 | 23 | 8.2 | 0.217 |
| *Mycophenolate mofetil* | 28 | 8.7 | 24 | 8.6 | 0.948 |
| *Tocilizumab* | 14 | 4.4 | 23 | 8.2 | 0.050 |
| *Rituximab* | 5 | 1.6 | 6 | 2.1 | 0.762* |
| *IL-17i* | 8 | 2.5 | 2 | 0.7 | 0.115* |
| *JAKi* | 8 | 2.5 | 4 | 1.4 | 0.396* |
| *Abatacept* | 4 | 1.2 | 2 | 0.7 | 0.690* |
| *Belimumab* | 3 | 0.9 | 3 | 1.1 | 0.591* |
| *Cyclophosphamide (pulse therapy)* | 3 | 0.9 | 3 | 1.1 | 1.000* |
| *Sulfasalazine* | 3 | 0.9 | 6 | 2.1 | 0.316* |

Chi square test, unless otherwise noted. \*Fisher exact test; \*\*Mann-Whitney test; CI 95%. SLE: systemic lupus erythematosus; RA:
rheumatoid arthritis; Axial SpA: axial spondyloarthritis; Pso arthritis: psoriatic arthritis; TNFi: tumour necrosis factor inhibitor; IL-17i:
interleucine 17 inhibitor; JAKi: janus kinase inhibitor

In the group of patients with COVID-19 (n=321), the most frequent symptoms were headache (60.6%), fever (55.6%), dysgeusia (54.3%), asthenia (53.7%), anosmia (52.5%) and cough (49.1%). Dyspnea was reported in 35.1% of cases. The medications used to treat COVID-19 were analgesics (53.7%), azithromycin (40.7%), oral corticosteroids (20.2%) and hydroxychloroquine (10.2%), but 16.1% did not use any medication.

Regarding COVID-19 outcomes, hospital care was sought by 77 (23.9%) patients and 29 (9.0%) were hospitalized. Of these, 3 patients were admitted to the intensive care unit and used mechanical ventilation. None of the patients died.

### Disease activity assessment

There was no significant difference comparing cases and controls in a 6-month follow-up (V1, V2, and V3), regardless of IMRD (Figure 1).

**Figure 1.**
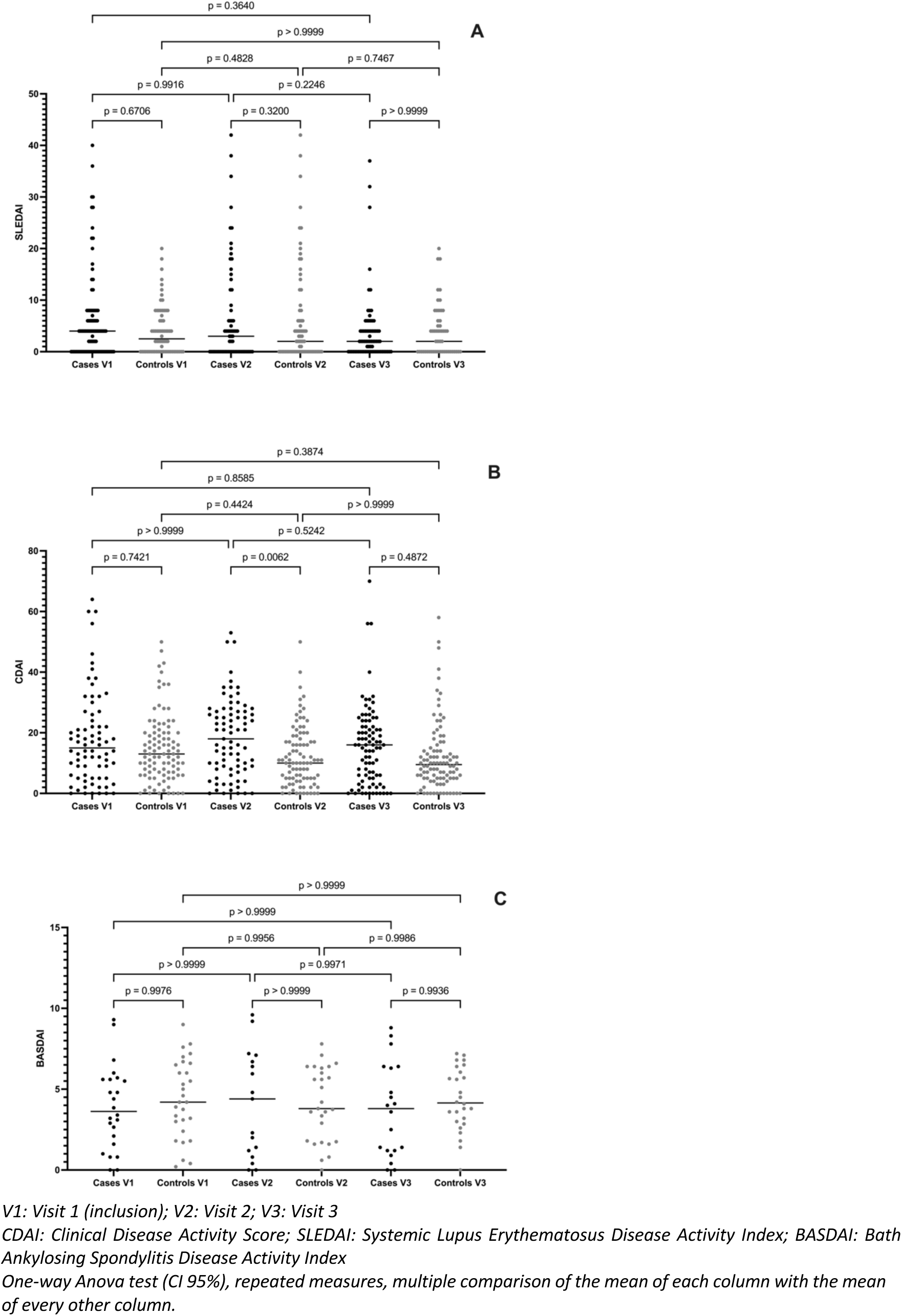
Comparison of disease activity scores across the three study visits (v1, V2 e V3) in patients with systemic lupus erythematosus, measured by SLEDAI (A), rheumatoid arthritis, measured by CDAI (B) and axial spondylarthritis patients, measured by BASDAI, comparing cases and controls matched to sex, age and epidemiological exposition.

Although no difference was demonstrated in the mean activity scores, some patients self-reported worsening of the disease activity after COVID-19 [30 with RA (32.2%), 32 with SLE (23.3%) and 2 with SpA (8.6%)]. Exploring these groups further, we also did not observe any significant difference in the mean activity score in RA and SLE patients (Figure 2).

**Figure 2.**
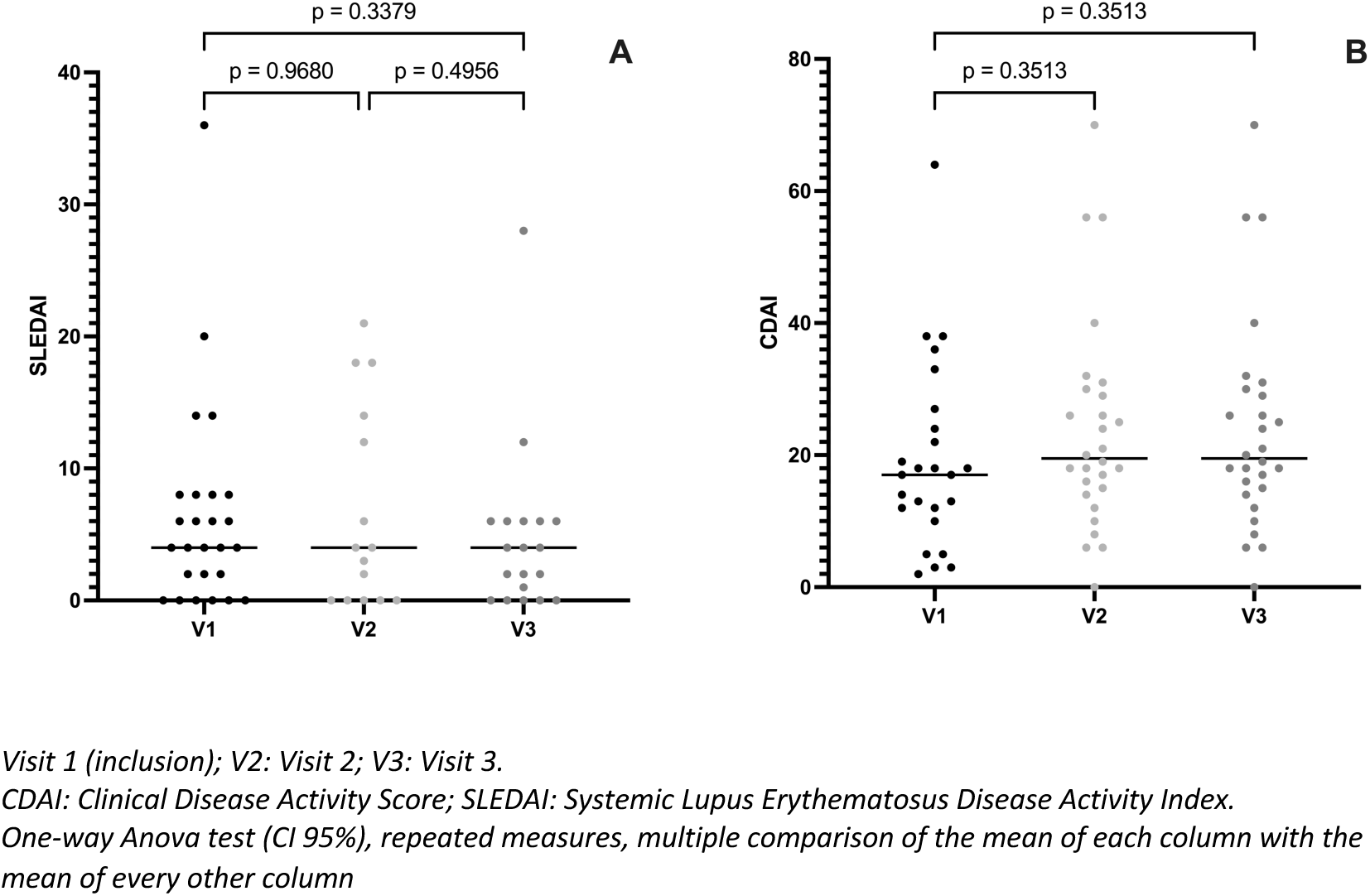
Comparison of disease activity scores across the three study visits, only in patients with systemic lupus erythematosus (SLEDAI) (A) and rheumatoid arthritis (CDAI) (B) who self-reported clinical worsening after COVID-19

In RA patients that self-reported worsening of disease activity after COVID-19, we applied the flare definition, comparing the CDAI pre-COVID with the CDAI post-COVID (inclusion), visit 2 and visit 3, in a paired way. At inclusion, 12 patients (40.0%) had an increased CDAI ≥ 4.5 points compared to pre-COVID status, but only 1 required a change in treatment. At visit 2, compared with visit 1, 5 new patients presented an increased CDAI ≥ 4.5 points and 7 patients improved compared to the previous visit. At visit 3, 12 patients presented an increased CDAI, and of these, 8 new patients, who had not worsened at visit 2, and only one needed change of treatment.

Besides, we compared the tender joint count (TJC), swollen joint count (SJC), patient global assessment (PGA) and medical global assessment (MGA) across the three visits, no significant difference was observed. Eight patients in this group (26.6%) self-reported worsening of joint manifestations.

Comparing the disease activity in the group of RA patients who self-reported worsening with those who did not worse, there was an association with post-COVID MGA, assessed at baseline (ES= – 0.56; CI95% –1.01 to –0.11, p=0.007) and with CDAI and all its components at visit 3 (Table 2).

**Table 2.**
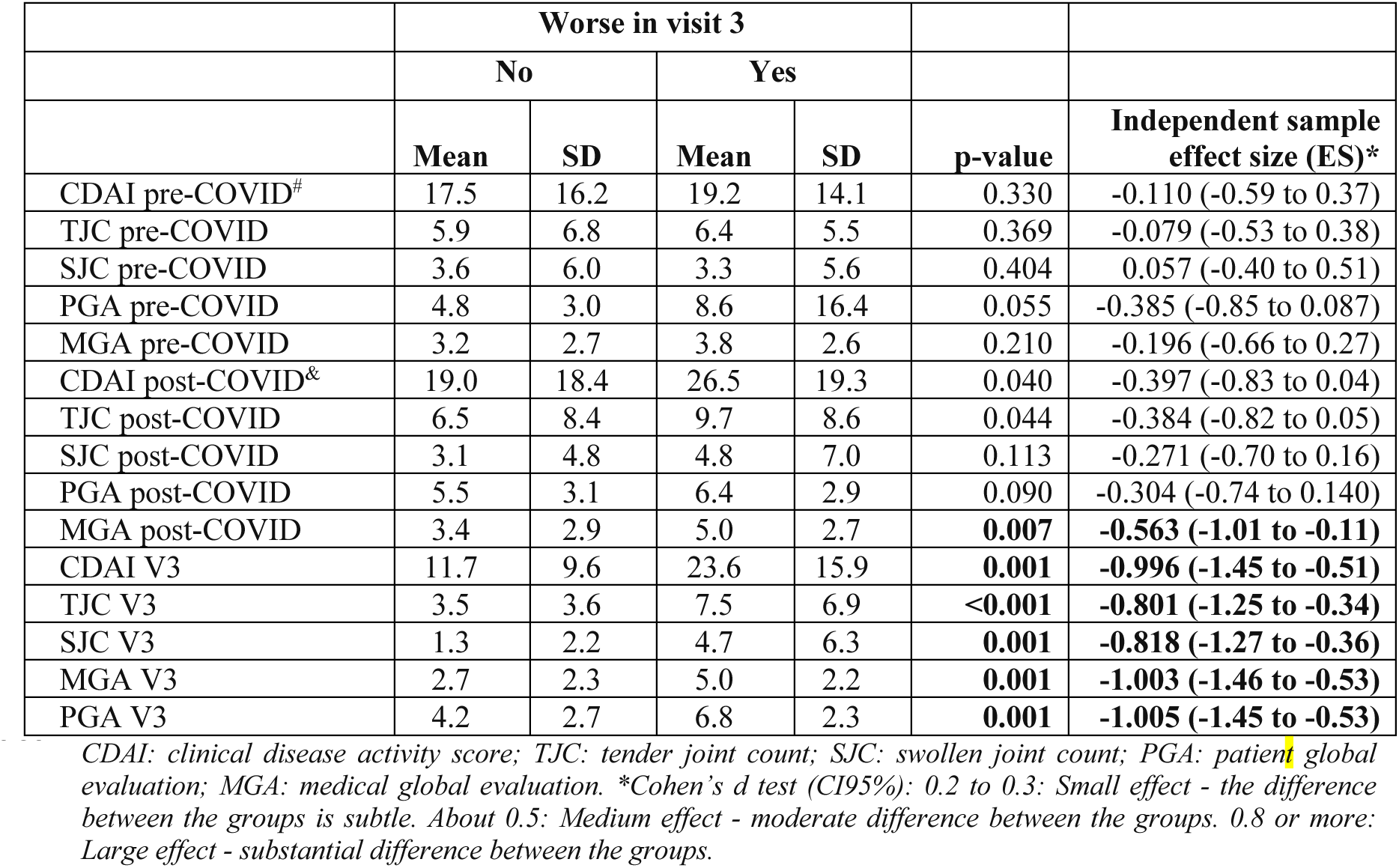
Comparison between Pre– and Post-COVID mean scores in patients with RA who reported worsening versus those who did not report it in the visit 3.

|  | Worse in visit 3 |  |  |  |  |  |
| --- | --- | --- | --- | --- | --- | --- |
|  | No |  | Yes |  |  |  |
|  | Mean | SD | Mean | SD | p-value | Independent sample effect size (ES)* |
| CDAI pre-COVID<sup>#</sup> | 17.5 | 16.2 | 19.2 | 14.1 | 0.330 | -0.110 (-0.59 to 0.37) |
| TJC pre-COVID | 5.9 | 6.8 | 6.4 | 5.5 | 0.369 | -0.079 (-0.53 to 0.38) |
| SJC pre-COVID | 3.6 | 6.0 | 3.3 | 5.6 | 0.404 | 0.057 (-0.40 to 0.51) |
| PGA pre-COVID | 4.8 | 3.0 | 8.6 | 16.4 | 0.055 | -0.385 (-0.85 to 0.087) |
| MGA pre-COVID | 3.2 | 2.7 | 3.8 | 2.6 | 0.210 | -0.196 (-0.66 to 0.27) |
| CDAI post-COVID<sup>&</sup> | 19.0 | 18.4 | 26.5 | 19.3 | 0.040 | -0.397 (-0.83 to 0.04) |
| TJC post-COVID | 6.5 | 8.4 | 9.7 | 8.6 | 0.044 | -0.384 (-0.82 to 0.05) |
| SJC post-COVID | 3.1 | 4.8 | 4.8 | 7.0 | 0.113 | -0.271 (-0.70 to 0.16) |
| PGA post-COVID | 5.5 | 3.1 | 6.4 | 2.9 | 0.090 | -0.304 (-0.74 to 0.140) |
| MGA post-COVID | 3.4 | 2.9 | 5.0 | 2.7 | **0.007** | **-0.563 (-1.01 to -0.11)** |
| CDAI V3 | 11.7 | 9.6 | 23.6 | 15.9 | **0.001** | **-0.996 (-1.45 to -0.51)** |
| TJC V3 | 3.5 | 3.6 | 7.5 | 6.9 | **<0.001** | **-0.801 (-1.25 to -0.34)** |
| SJC V3 | 1.3 | 2.2 | 4.7 | 6.3 | **0.001** | **-0.818 (-1.27 to -0.36)** |
| MGA V3 | 2.7 | 2.3 | 5.0 | 2.2 | **0.001** | **-1.003 (-1.46 to -0.53)** |
| PGA V3 | 4.2 | 2.7 | 6.8 | 2.3 | **0.001** | **-1.005 (-1.45 to -0.53)** |

CDAI: clinical disease activity score; TJC: tender joint count; SJC: swollen joint count; PGA: patient global evaluation; MGA: medical global evaluation. \*Cohen's d test (CI95%): 0.2 to 0.3: Small effect - the difference between the groups is subtle. About 0.5: Medium effect - moderate difference between the groups. 0.8 or more: Large effect - substantial difference between the groups.

The size effect for MGA post COVID (V1) mean difference means was moderate (–0.56; CI95% –1.01 to –0.11) and for CDAI (–0.996; CI95% –1.45 to –0.51), TJC (–0.801; CI 95% –1.25 to –0.34), SJC (–0.818; CI-1.27 to –0.36), MGA (–1.003; CI –1.46 to –0.53) and PGA at V3 (–1.005; CI –1.45 to –0.53) was a large effect, demonstrating substantial difference between the groups.

Post-COVID worsening in patients with RA was associated with diabetes mellitus (OR=7.15; CI95% 1.7-29.4, p=0.005) and protectively with current use of TNF inhibitors before the SARS-CoV-2 infection (OR=0.51; CI95% 0.2-0.9, p=0.026).

Among the patients with SLE who self-reported worsening of disease after COVID-19, 10 of them reported the appearance of new clinical manifestations. However, no significant differences were observed regarding gender, age, comorbidities, or concomitant medications. Worsening was only associated with anosmia, as a symptom of COVID-19 (OR=2.43; CI95% 1.07-5.52, p=0.03). The self-reported clinical manifestations by SLE patients who were worse after COVID-19 (n=32) at visit 3 were skin rash (5 patients), arthritis (3 patients), proteinuria (2 patients) and interstitial lung disease (1 patient).

Comparing the SLEDAI pre– and post-COVID through the flare definition, we observed that 6 patients (18.0%) had an increased SLEDAI ≥ 4.0 points, but only 2 required a change in treatment. At visit 2, compared with visit 1, most patients showed SLEDAI improvement and only 3 patients had it increased at visit 3. However, 3 new patients had an SLEDAI increasing compared with the visit 2.

Among SpA patients, only 2 patients self-reported worsening after COVID-19 and one of them had progressive worsening in all three visits, based on the BASDAI.

There was no statistically significant association between COVID-19 outcomes (hospital care, hospitalization, and ICU admission) with post-COVID disease activity at visit 3.

### Fatigue, depression, anxiety, and stress assessment

After a 6-month follow-up, a statistically significant difference was observed through FACIT and DASS 21 questionnaires in their three domains comparing cases and controls (Figure 3).

**Figure 3.**
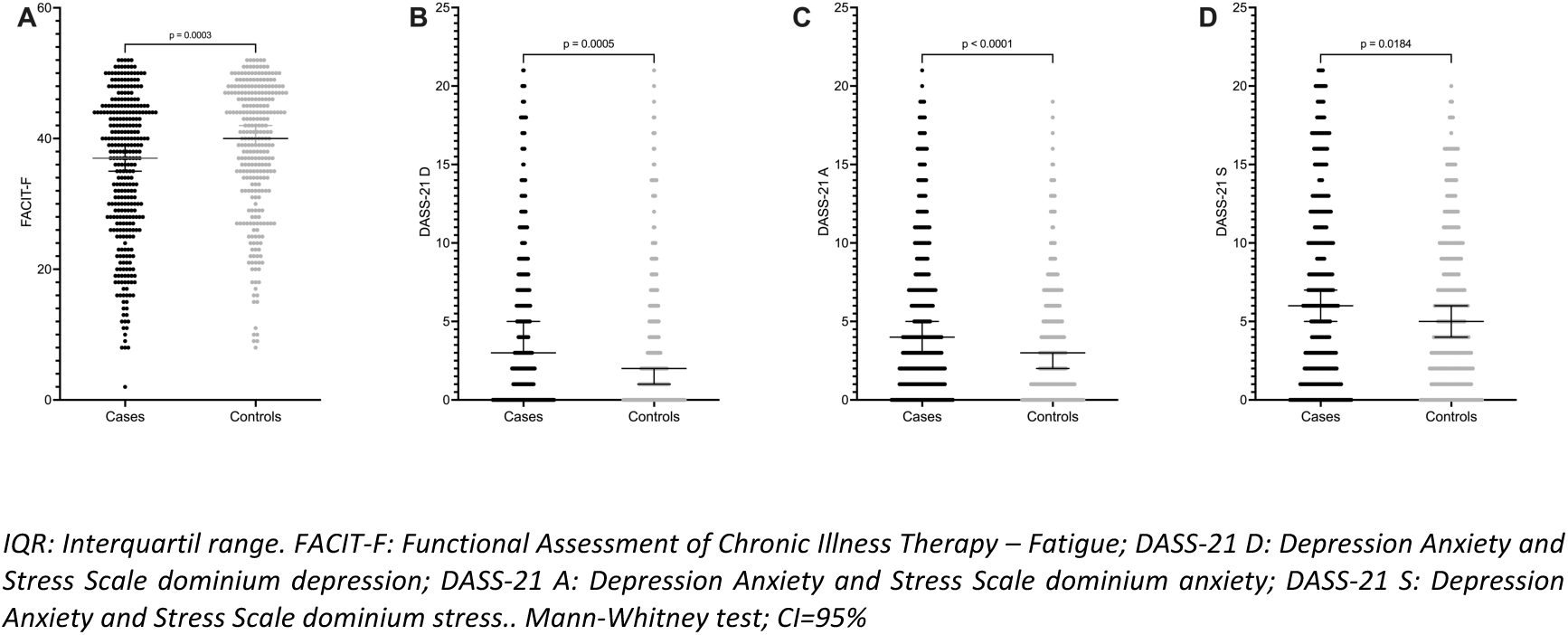
Comparison among FACIT (A), DASS-21 D (B), DASS-21 A (C) and DASS-21 S (D) scores median (IQR) in cases and controls at visit 3.

The median FACIT score was 37 (27–44) in cases and 40 (32–47) in controls (p=0.0003); DASS-21 D median in cases was 3 (0-8) and 2 (0-5) in controls (p=0.0005); DASS-21 A median in cases was 4 (1–9) and 3 (1–6) in controls (p<0.001) and DASS-21 S median in cases was 6 (2-12) and 5 (2-9) in controls (p<0.018) No difference was observed regarding the FACIT-F and the DASS-21 scores when comparing patients with RA, SLE and SpA in each case group (Figure 4).

**Figure 4.**
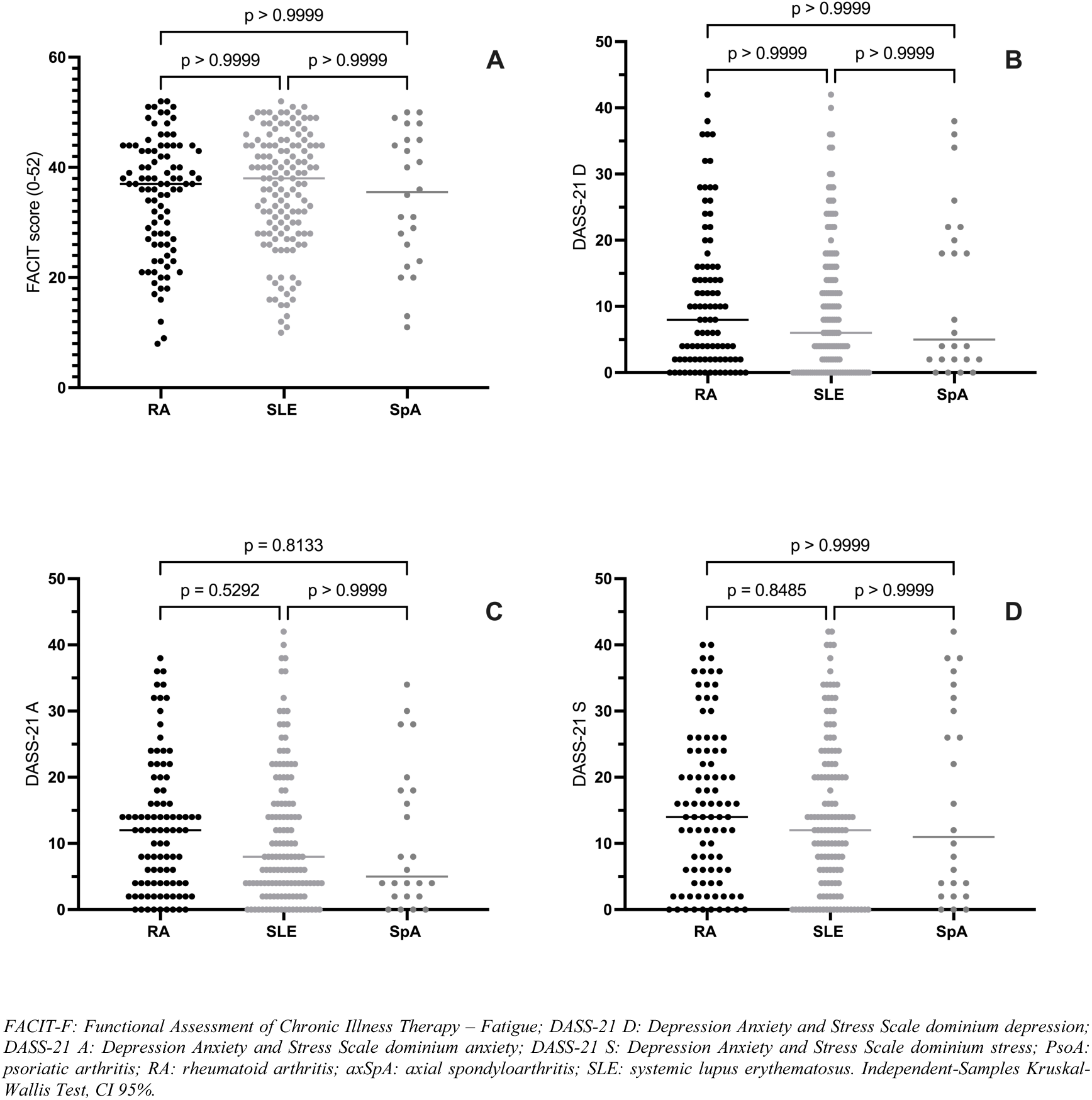
Distribution and comparison among FACIT (A), DASS21 D (B), DASS 21 A (C) and DASS 21 S (D) median scores after a 6-month follow-up in patients with rheumatoid arthritis (RA), systemic lupus erythematosus (SLE) and axial spondyloarthritis (axSpA) only in cases group (IMRD COVID-19 +)

There was no association of FACIT or DASS-21 with comorbidities, IMRD endpoints (type and specific therapy), and outcomes related to COVID, such as symptoms and treatment, in univariate linear models. Considering the FACIT-F and DASS-21 scores, there was not significant correlation between the disease activity in RA and SLE patients from case group (Figure 5).

**Figure 5.**
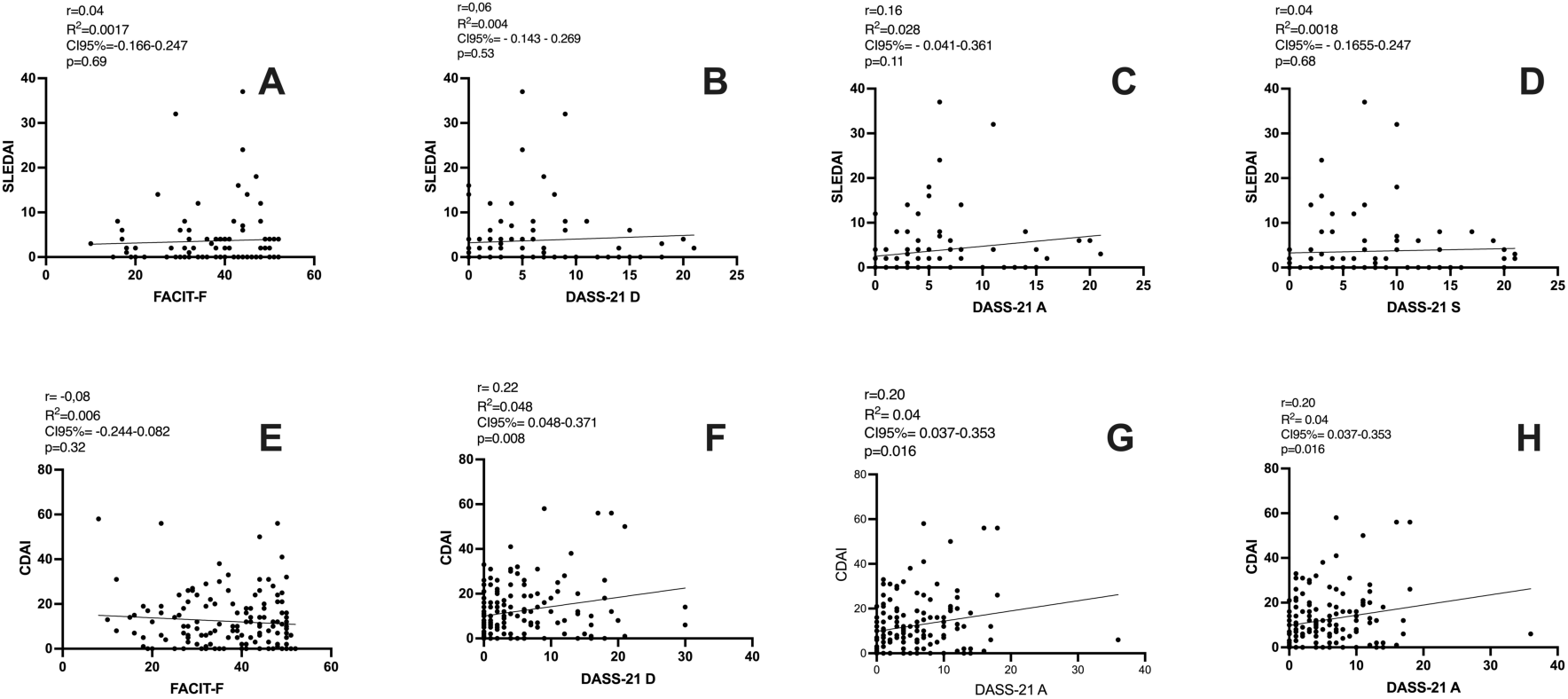
Correlation between disease activity in patients with systemic lupus erythematosus, measured by SLEDAI, and rheumatoid arthritis, measured by CDAI, only in case group (IMRD COVID-19 +), with FACIT-F (A and E) and DASS 21-D (B and F), DASS 21-A (C and G) and DASS 21-S (D and H) scores

Specifically analyzing the group of patients with worsening of disease activity after COVID-19, we observed that RA patients also had a worsening of FACIT-F (31.5 vs. 36.4; p=0.047) and DASS-21-S (19.0 vs. 13.0; p=0.031) when compared to those witn no worsening activity. Also, the means was considered as moderate for FACIT<u>-F</u> (0.47; from 0.03 to 0.91) and DASS 21-S (–0.44; from –0.88 to –0.007) (Table 3). In SLE patients, no significant difference was observed.

**Table 3.**
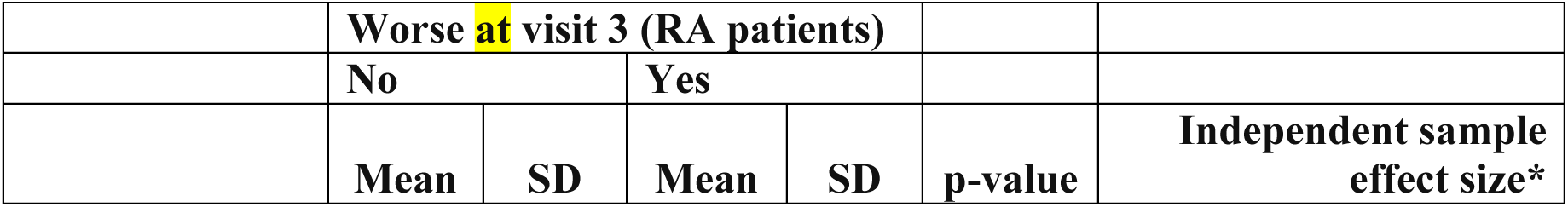
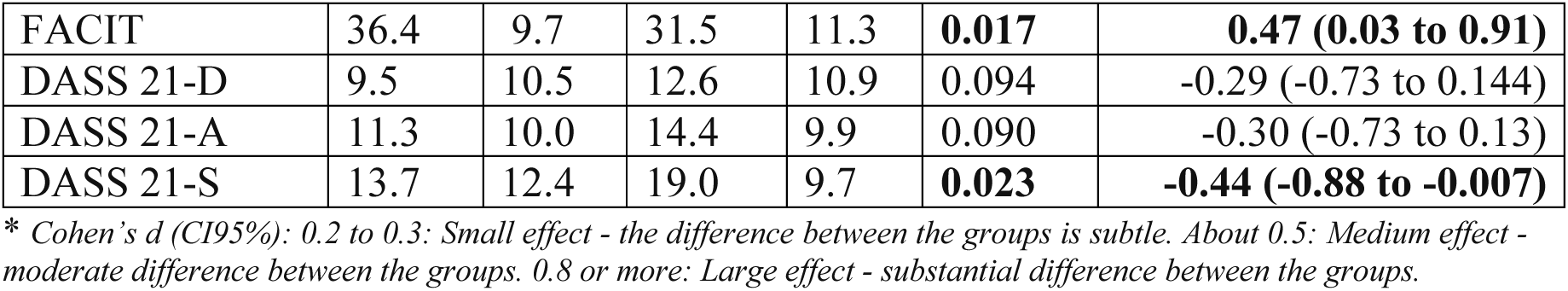
Comparison among mean scores for FACIT, DASS 21-D, DASS 21-A and DASS-21 S in patients with RA who reported worsening versus those who did not report it.

There was no statistically significant association between COVID-19 outcomes (hospital care, hospitalization, and ICU admission) with FACIT-F and DASS-21 scores at visit 3.

## Discussion

Our results demonstrated that the disease activity scores did not differ significantly in IMRD patients during a 6-month follow-up after COVID-19 in comparison to those who did not contract the virus. However, the clinical worsening was related to fatigue, depression, anxiety, and stress.

Nowadays, the data regarding the association between SARS-CoV-2 infection and IMRD flare are controversial, especially because the published studies have heterogeneous samples and different designs. Consequently, the current uncertainty revolves around whether the Patient-Reported Outcomes (PROs) are related to IMRD flare itself after COVID-19 or if it is associated with Long Covid or, alternatively, represent a reactive post-infectious immunological response working a potential trigger (1, 12–15).

In rheumatic diseases, a flare is defined as any exacerbation of disease activity that, if persistent, would typically necessitate the initiation or modification of therapy. It signifies a cluster of symptoms of sufficient duration and intensity to warrant the commencement, alteration, or escalation of therapeutic measures (16). In our study, although there was no discernible difference in mean activity scores among patients with rheumatoid arthritis (RA), systemic lupus erythematosus (SLE), and spondyloarthritis (SpA), a subgroup of patients reported a deterioration in immune-mediated rheumatic disease (IMRD). In some of these cases, activity scores increased, aligning with the flare definition employed in this investigation. Given the recognized potential for overestimation of IMRD activity tools in patients with conditions like fibromyalgia (17) and other chronic painful disorders, particularly in RA and axial SpA, it becomes imperative to ascertain the presence of “true” disease activity. This determination holds substantial relevance in decision-making processes related to treatment adjustments. Nevertheless, we observed an association between patients’ self-reported worsening after 6 months and the Clinical Disease Activity Index (CDAI) score, along with all its individual components, including swollen joint count (SJC). This association suggests a plausible occurrence of flare in this specific subset of patients.

A cross-sectional study, including 32 patients with RA and SpA, demonstrated no relevant change regarding disease activity after COVID-19 in those who interrupted their treatment (1). Data from the COVAD study, a cohort that evaluated 824 patients with IMRD who had at least one SARS-CoV-2 infection, between March 2021 and June 18, 2022 (12), showed that 36.9% of patients experienced at least one flare of the underlying IMRD following COVID-19 infection over a 127 day period (IQR = 62-308 days) from the date of infection to the date of the survey. Females and patients with comorbidities had higher odds of flaring of their disease. Patients who reported flares had worse physical health scores, pain VAS, fatigue VAS, and lower mental health scores compared to those who did not report flares. However, as stated by the authors, “this was a self-reported disease flares without verification by a physician which could be impacted by patients’ perceptions of flares and their inability to distinguish from ongoing symptoms of long-COVID syndrome or secondary fibromyalgia”.

In a study conducted on 92 children with IMRD who have had COVID-19 (13), 10% had a relapse of IMRD after infection. The relapse was mild in four and moderate in five cases. One patient had a severe relapse of ARD and required hospitalization, without association with COVID-19 clinical presentation.

Similar to other several viral infections, SARS-CoV-2 infection could be potential trigger of reactive and autoimmune diseases by inducing type II and type IV hypersensitivity reactions, leading to autoantibodies production and autoimmune diseases development as long-term complications (18), and in patients with pre-existing IMRD, it may be difficult to identify whether it is a flare of the disease or a post-infection manifestation. A recent study evaluated the effects of COVID-19 on the development and progression of RA using a collagen-induced arthritis (CIA) animal model. The incidence and severity of RA in CIA mice were slightly increased by SARS-CoV-2 spike protein in vivo. In addition, the levels of autoantibodies and thrombotic factors, such as anti-CXC chemokine ligand 4 (CXCL4, also called PF4) antibodies and anti-phospholipid antibodies were significantly increased by SARS-CoV-2 spike protein. Furthermore, tissue destruction and inflammatory cytokine level in joint tissue were markedly increased in CIA mice by SARS-CoV-2 spike protein, suggesting that COVID-19 accelerates the development and progression of RA by increasing inflammation, autoantibody production, and thrombosis(19).

In an Italian study, 122 consecutive post-COVID-19 cases were evaluated, with the onset of the rheumatic manifestations within 4 weeks from SARS-CoV-2 infection as an inclusion criterion. In this group, it was identified that most patients had inflammatory joint disease (52.5%); 19.7% of cases were diagnosed with connective tissue diseases and 6.6% of cases with vasculitis. It is interesting to note that in this same cohort, patients with inflammatory manifestations post-COVID-19 vaccine were evaluated, and this group had a higher prevalence of patients classified as polymyalgia rheumatica (PMR, 33.1% vs. 21.3%, p=0.032) (14). Another study evaluated the presence of arthritis associated with COVID-19 by ultrasound in 10 patients with (n=4) and without previous rheumatic disease (n=6). In the group without previous disease, 4 of the 6 patients presented arthritis for 4 to 16 weeks after infection, comparable to reactive arthritis. One of them developed late-onset rheumatoid arthritis. In the group with previous disease, synovitis and tenosynovitis with positive power Doppler were observed, suggesting a possible flare of the disease after COVID-19 (20).

A major concern when interpreting the musculoskeletal symptoms in these patients is the possibility of developing post-Covid syndrome, also known as long Covid. More than 20% of subjects surviving acute COVID-19 may suffer from persisting symptoms and develop new ones after one month and about 5% of all infected individuals develop long-term complications after 6 months, possible due to tissue damage, viral reservoirs, autoimmunity, and persistent inflammation (21, 22). The clinical presentation in these cases, such as fatigue and joint pain, may mimic a disease flare, thereby complicating the decision-making regarding treatment’s change.

Recent systematic review showed that the prevalence of arthralgia ranges from 2% to 65% within a time frame varying from 4 weeks to 12 months after COVID-19. Inflammatory arthritis has been reported with various clinical phenotypes, including RA-like pattern as other prototypical viral arthritis, as well as polymyalgia-like, or acute monoarthritis and oligoarthritis of large joints resembling reactive arthritis or SpA style (23).

An Iranian study evaluated the prevalence of musculoskeletal symptoms in 239 patients after the acute phase of COVID-19 and its associated factors, using an online questionnaire. Almost all of them (98.74%) had experienced at least one musculoskeletal symptom after recovering from COVID-19, and the most common symptom was fatigue (91.2%), followed by myalgia, headache, and low back pain. High BMI, hospitalization, and ICU admission were associated with a higher risk of musculoskeletal symptoms (24).

Besides musculoskeletal symptoms, patients could develop psychological distress after COVID-19, such as depression, anxiety, and stress, symptoms that could also be present in IMRD. In our patients, the frequency of these symptoms in IMRD post COVID-19 was significantly higher than in patients who did not have the infection. Some authors suggested that the term of long COVID is unappropriated and should be replaced by fibromyalgia-like post-COVID syndrome (2).

We acknowledge the limitations of our study, such as the fact we have the FACIT and DASS 21 scores only at the last visit and no possibility of comparison with the pre-COVID status. Since the study began in the first months of the pandemic, we did not know yet that these outcomes were important. We decided to include it after the first studies were published demonstrating the association of these symptoms with SARS-CoV-2 infection. However, we know that fatigue, depression, anxiety, and stress are frequent symptoms in patients with IMRD and we have a group that did not have COVID-19 as a comparator, and in the group with COVID-19 these scores were significantly worse. In addition, the greater political and economic instability experienced in our country may have had a direct impact on psychological distress as a bias. On the other hand, we have a representative sample, from various regions of Brazil, with a follow-up period of 6 months, compared with a group of patients with IMRD matched by sex and age, for the same epidemiological period, one of the main strengths of our study.

## Conclusions

Our observational data underscore the complex interplay between COVID-19 and immune-mediated rheumatic diseases (IMRD). While disease activity scores may not exhibit significant differences, individuals with post-COVID-19 IMRD may undergo noteworthy deterioration in psychological well-being. The variations in reports regarding IMRD flares and the potential triggering role of SARS-CoV-2 in autoimmune manifestations underscore the need for thorough clinical assessment and a comprehensive approach to their management.

## Data Availability

All data produced in the present study are available upon reasonable request to the authors

## Acknowledgements

All ReumaCoV Brazil researchers

